# Innovative Approaches in Pericardiocentesis Training: A Comparative Study of 3D-Printed Mannequins and Virtual Reality Simulations

**DOI:** 10.1101/2024.02.16.24302932

**Authors:** Alberto Rubio-López, Rodrigo García-Carmona, Laura Zarandieta-Román, Alejandro Rubio-Navas, Ángel González-Pinto, Pablo Cardinal-Fernández

**Affiliations:** Head of Intensive Care Unit. HM Monteprincipe University Hospital. Madrid. Spain; Universidad CEU San Pablo School of Medicine, Madrid, Spain; Department of Computer Science. Universidad CEU San Pablo. Madrid. Spain; Biomedical Engineer. Universidad CEU San Pablo. Madrid. Spain; Biology Graduate. Universidad Autónoma. Madrid. Spain; Head of Cardiac Surgery Unit. Gregorio Marañón University Hospital. Madrid. Spain; Head of Cardiac Surgery Unit. HM Monteprincipe University Hospital. Madrid. Spain; Head of Intensive Care Unit. HM Torrelodones University Hospital. Madrid. Spain

**Keywords:** Intensive Care training, Pericardiocentesis, Simulation models, 3D-printed mannequin, Virtual reality (VR) training, Medical education, Heart rate variability (HRV), Stress analysis in medical training

## Abstract

**Background:** The CoBaTrICE initiative standardized intensive care training throughout Europe. Pericardiocentesis, an important yet infrequent operation, poses unique training challenges often unmet by conventional methods. This study evaluates the efficacy of virtual reality (VR) simulations and 3D-printed mannequins in this procedure training, both crafted from scratch using highly affordable materials and free software.

**Methods:** Thirty-five final-year medical students participated in this prospective, quasi-experimental study. Initially, students underwent VR simulation training, followed by training using 3D-printed self-made mannequins. Learning outcomes were assessed using the Objective and Structured Clinical Examination (OSCE) questionnaire. Heart rate variability (HRV) analysis was employed to monitor stress responses, while the NASA Task Load Index (NASA-TLX) was used to gauge self-perceived difficulty.

**Results:** Although the mannequin model induced more stress reactions, it excelled in tasks requiring fine motor skills, such as correct placement and asepsis maintenance. In contrast, lower NASA-TLX scores indicated that the VR model imposed less mental demand and effort, reflecting a reduced cognitive load.

**Conclusion:** Both VR simulations and 3D-printed mannequins effectively taught pericardiocentesis skills, each offering distinct advantages. Mannequins enhanced fine motor skills, whereas VR reduced cognitive load and increased engagement. The combination of both approaches could maximize training outcomes, particularly in resource-constrained environments, thus broadening access to advanced simulation education.

## Introduction

The CoBaTrICE initiative, established in 2003, has played a crucial role in standardizing intensive care medicine education across Europe[1]. This program provides a flexible framework for advanced medical training, including the critical yet infrequent procedure of pericardiocentesis [2]. Conventional teaching methods often fall short in adequately preparing physicians for this invasive and rarely performed technique, presenting a need for innovative training solutions [3].

Clinical simulation has emerged as a valuable tool in addressing these training challenges. These simulations range from simple models to sophisticated digital systems, following Kolb’s Experiential Learning Cycle [4], which emphasizes tangible experience, reflective observation, abstract conceptualization, and active experimentation. Despite their benefits [5], high-fidelity simulators are costly and require specialized infrastructure and highly knowledgeable instructors, posing challenges for resource-limited universities [6].

Recently, 3D printing technology has been integrated into medical education, allowing the creation of realistic anatomical models at a reduced cost. This innovative approach enables students to engage in interactive, hands-on learning environments, enhancing the accessibility and quality of instructional materials [7].

Virtual reality (VR) represents another transformative technology in medical training [8]. VR offers numerous advantages over traditional simulation techniques, including increased participation, precise replication, consistent training scenarios, emotional engagement, and future development potential [9]. With cost-effective programming tools, final-year students can create diverse and sophisticated VR scenarios, offering a scalable and affordable training solution [10].

The effectiveness of these training methods is often evaluated using the Objective Structured Clinical Examination (OSCE) [11], a standardized tool that assesses a broad range of clinical competencies [12]. Additionally, understanding the self-perceived difficulty of trainees through instruments like the NASA Task Load Index (NASA-TLX) is crucial for refining simulation-based training to meet learners’ needs effectively [13].

Nevertheless, there remains a significant gap in verifying the emotional impact of virtual reality (VR) simulations, especially regarding their ability to induce stress responses, which are critical in high-stakes medical procedures. This study aimed to evaluate the effectiveness of both traditional methods and VR simulations in eliciting authentic stress responses during pericardiocentesis, a procedure associated with considerable potential complications and heightened stress levels. We employed heart rate variability (HRV) analysis, a validated measure of stress [14], to assess and compare the effectiveness of a 3D-printed scenario and a VR simulation in preparing medical students for real-life challenges, intending to demonstrate VR’s potential in educational settings with limited resources.

This study had two primary objectives: first, to develop a 3D-printed mannequin and a VR simulation model for training using affordable tools and readily accessible software; second, to compare the learning outcomes, stress levels measured through biometric data, and self-perceived difficulty between these two simulation models. If both methods proved equally effective, this study would validate the use of virtual reality in complex medical procedures. This technology holds significant promise for medical institutions and universities, particularly those in low- and middle-income countries, due to its cost-effectiveness and the engagement of senior medical students.

## Materials and Methods

### Study design

This pilot prospective quasi-experimental study investigated two training models for pericardiocentesis: a self-built mannequin made using 3D design and printing techniques, and a Unity-based virtual reality (VR) environment.

### Creation and Validation of the Training Models

The mannequin model was developed using 3D modeling tools, specifically Tinkercad and Adobe Fusion 360, to create a computer model reflecting the anatomical elements relevant to this technique. The model was then 3D printed using Ultimaker Cura and MeshMixer (Figures 1-3). The VR model was developed using the Unity game engine to structurally match the physical model, ensuring consistency across training scenarios. The VR environment was compatible with HTC Vive, Oculus Rift, and Go systems (Figure 4).

**Figure 1.**
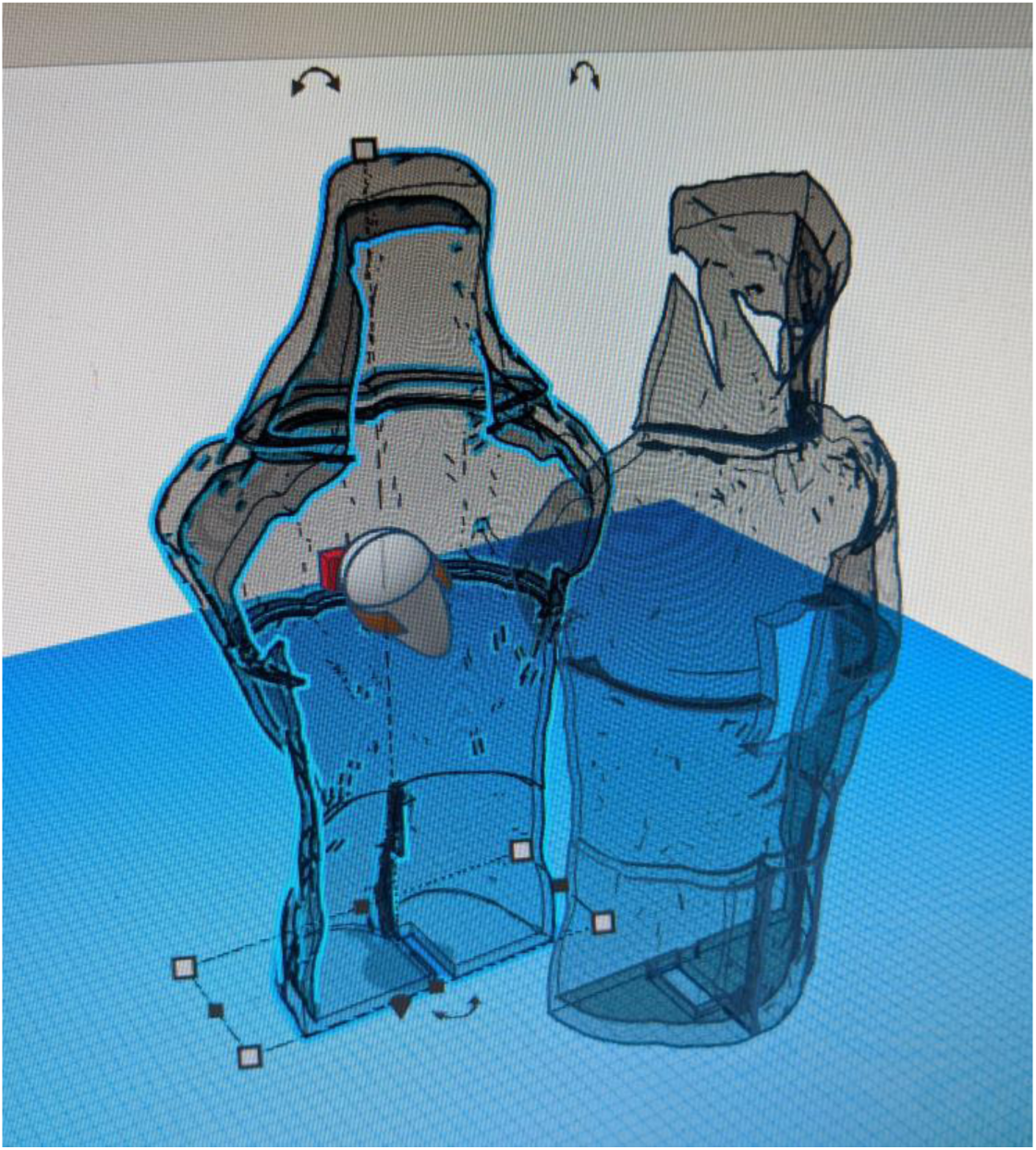
3D Design of mannequin, pericardium, and support

**Figure 2.**
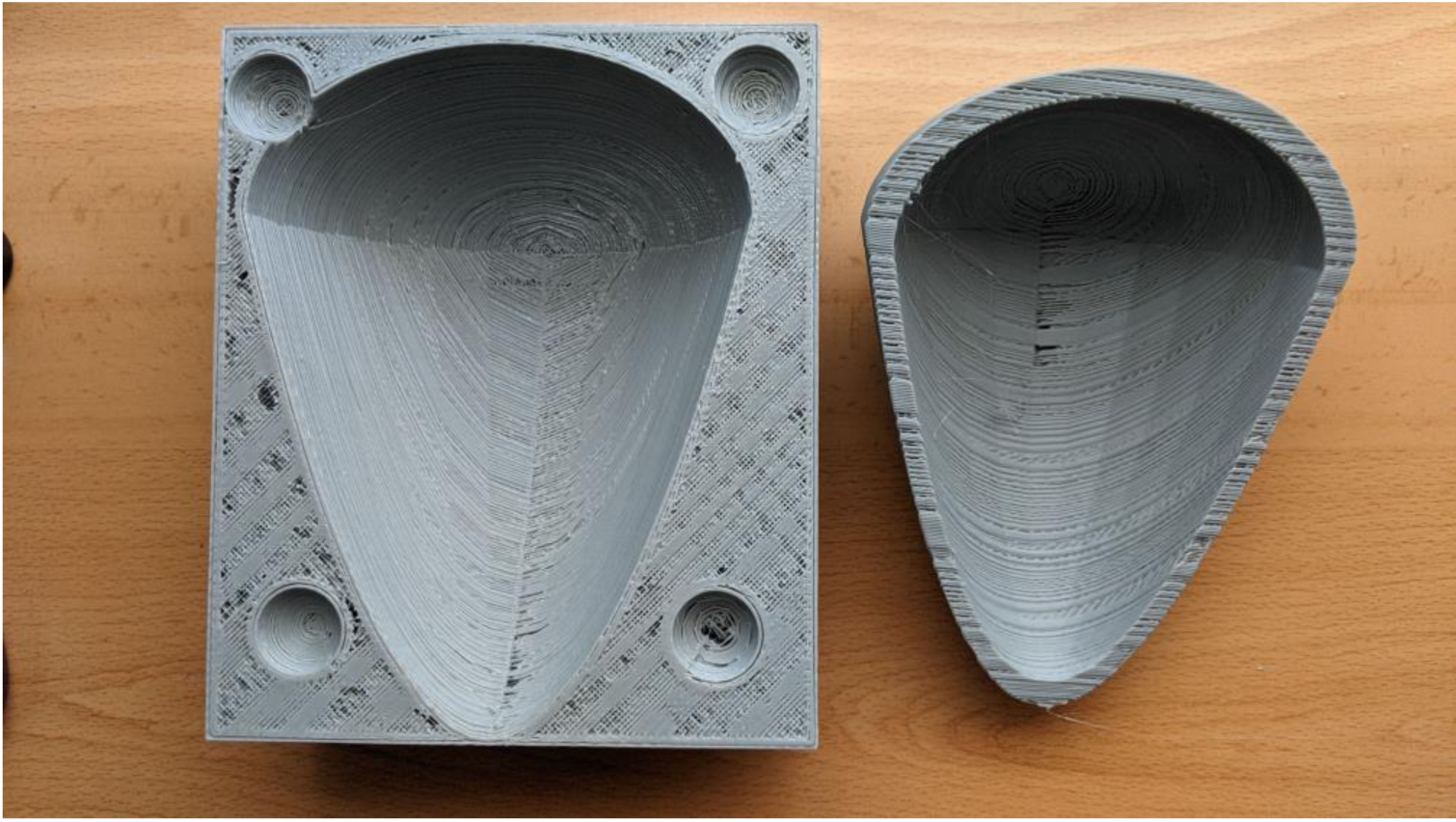
Pericardium 3D printed mold

**Figure 3.**
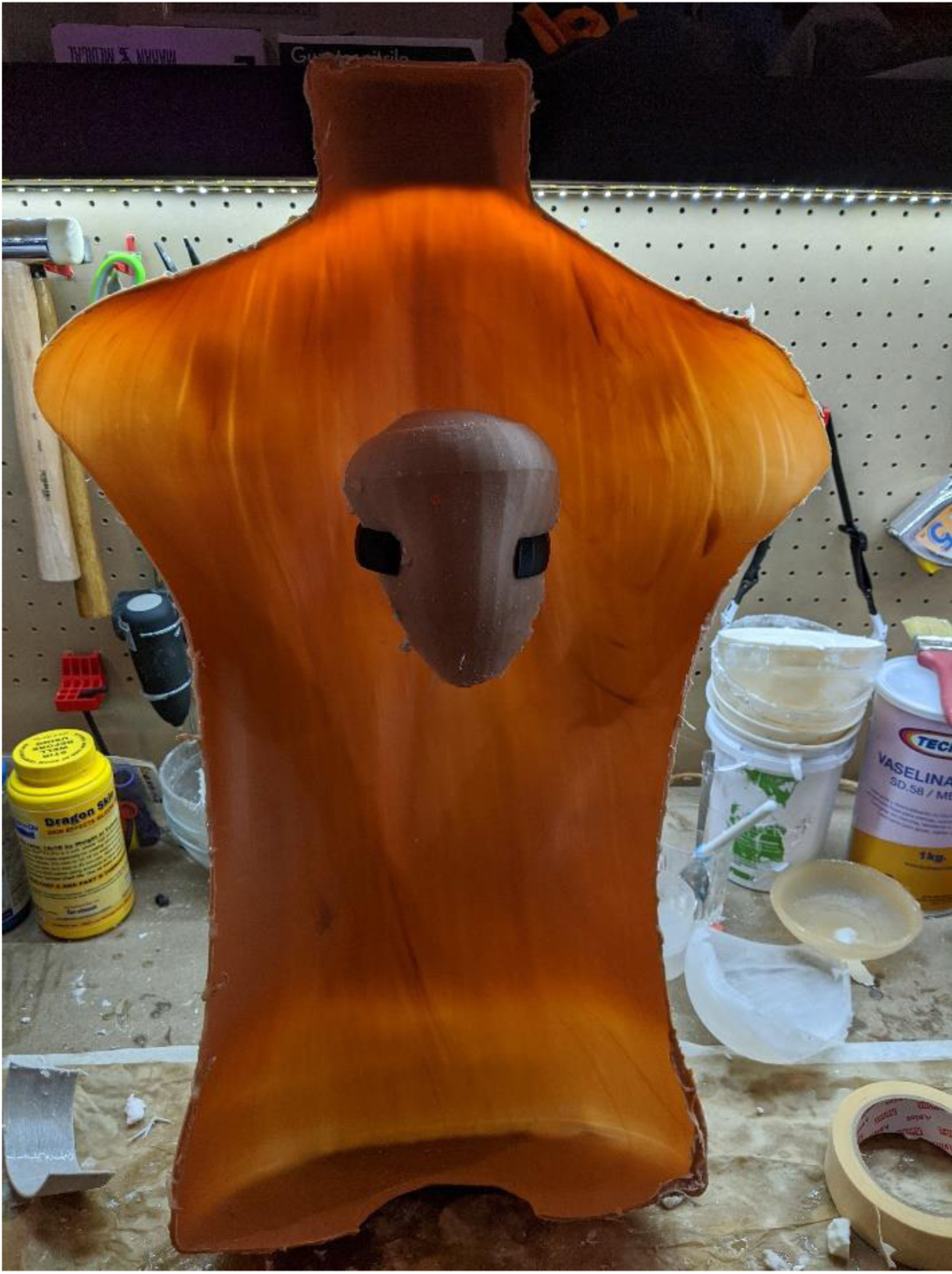
3D printed pericardium and support

**Figure 4.**
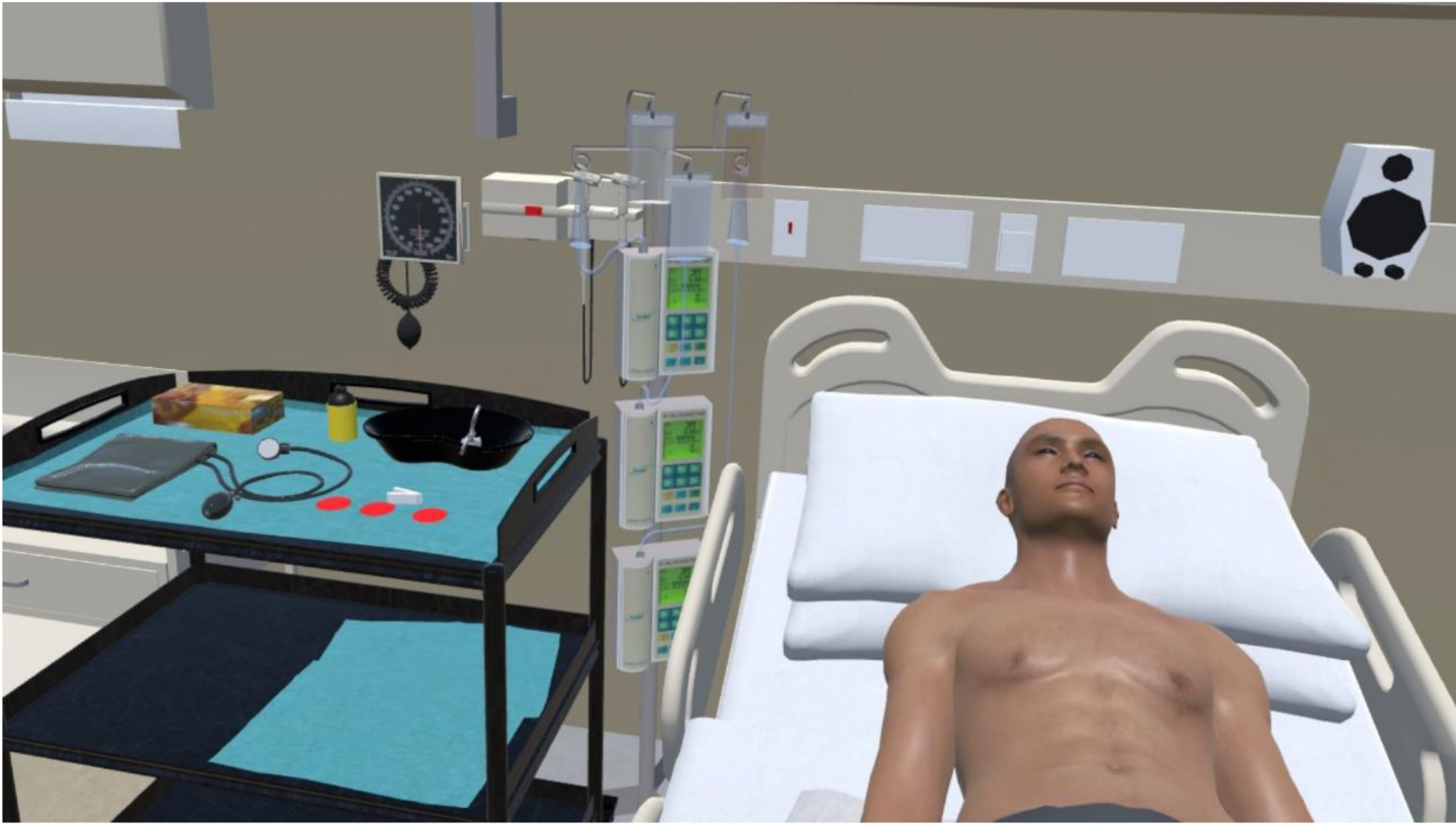
VR Scenario

Both models were fine-tuned and validated by five experienced intensivists from the Hospital’s Intensive Care Unit using a Delphi two-round process (Supplementary Materials)

### Ethical considerations and recruitment

The University Hospital Research Ethics Committee approved the study. Written informed consent was obtained from all participants, and their data were anonymized. Thirty-six final-year medical students were recruited for the study.

### Inclusion and Exclusion Criteria

All final-year medical students who provided written consent were included, except those on medication affecting heart rate. Participants with insufficient data or incorrect signals were excluded. All participants watched a standard teaching film on pericardiocentesis as part of the preparation (Appendix A).

### Randomization and Participant Grouping

Participants were evaluated by an external evaluator using the Objective Structured Clinical Examination (OSCE) Questionnaire (Supplementary Materials), which assessed diagnostic and procedural skills. Learning parameters included rapid response, blood pressure, cardiac rhythm analysis, pulse oximeter placement, sphygmomanometer use, material handling, glove placement, asepsis, surgical field placement, puncture site location, needle angulation, guidewire placement, and drain placement. Each item was scored on a scale from 0 to 5, with one global evaluation item.

To assess stress responses, prior to the initiation of the simulation, students were equipped with three electrodes on the torso for continuous heart rate recording via the biosignal plux system. Data acquisition and subsequent processing were executed using OpenSignals digital signal processing software (Figure 5 and Appendix B). Stress parameters were assessed according to heart rate variability (HRV) analysis, as it serves as a sensitive marker for dysregulation in the Autonomic Nervous System (ANS) and is defined as the temporal variation in the intervals between consecutive heartbeats over a predefined period.

**Figure 5.**
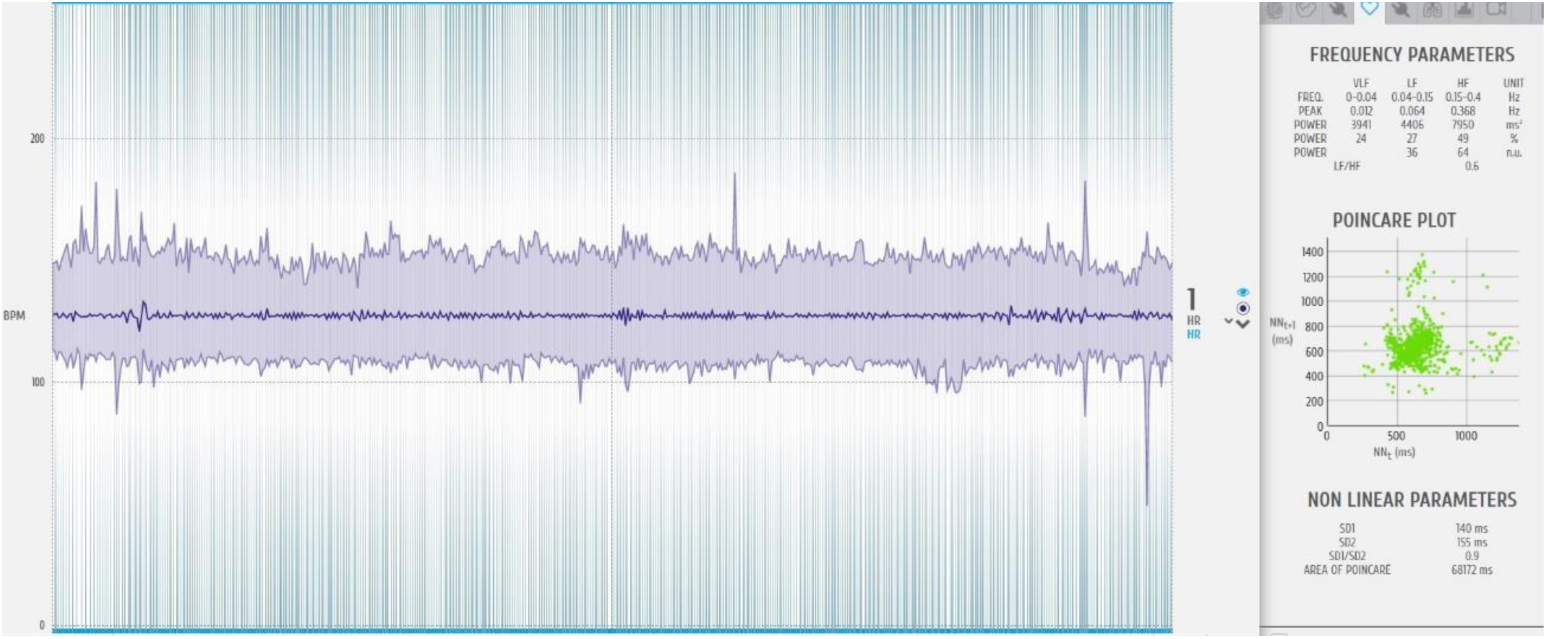
Biosignal plux digital signal processing tool

The commonly utilized parameters include frequency-domain parameters such as Low Frequency (LF), primarily associated with sympathetic activation but also including a parasympathetic component, High Frequency (HF), a specific indicator of parasympathetic nervous system (PNS) activity linked to stress relief, and the LF/HF Ratio, which serves as an indicator of sympathetic activity and is associated with heightened stress levels. Time-domain parameters include rMSSD, the root mean square of successive differences between normal heartbeats, closely related to the PNS, and PNN20 and PNN50, the percentage of differences associated with the number of intervals between heartbeats that vary by more than 20 and 50 milliseconds, respectively, both closely correlating with the PNS. Non-linear parameters, derived from the Poincaré plot, which is a visual representation of temporal intervals between consecutive heartbeats, include the SD1/SD2 Ratio, correlating with the LF/HF ratio.

These measurements allowed us to evaluate changes in stress parameters from the baseline state to the simulation scenarios and how each scenario (mannequin and VR) affects stress in comparison to the baseline state, enabling detection of any differences between the two simulation scenarios relative to the baseline state.

### Measurement of self-perceived difficulty

The NASA Task Load Index (NASA-TLX) was used to evaluate cognitive load across six subscales: Mental Demand, Physical Demand, Temporal Demand, Performance, Effort, and Frustration. After completing each scenario, participants rated their experience on these subscales to provide a comprehensive analysis of perceived workload.

### Data Collection Phases

Participants answered a questionnaire including demographic data, English competency level (needed to properly complete the NASA Task Load Index), past VR use, and health status before the study started. Baseline data collection involved compiling stress data while subjects rested, establishing a baseline for each participant.

The first training scenario involved students starting their instruction with virtual reality-based exercises, followed by a resting phase to allow students to return to their usual stress levels, thereby preparing for the second training session free from any aftereffects. In the second training scenario, participants used the mannequin model to compare learning outcomes and stress reactions between the two modalities.

### Statistical analysis

Using IBM SPSS version 25, we compared variables between the mannequin and VR groups to evaluate the effect of the training sequence on learning results, stress reactions, and self-perceived difficulty. A comparative study was conducted to investigate variations in learning, biometric stress and NASA-TLX factors among the two scenarios. Appropriate statistical tests were used for this analysis.

## Results

There was a total of thirty-five medical students who took part in the study. The mean age of the participants was 23.54 years, with a standard deviation of 1.69. The age range of the participants was from 22 to 30 years. The cohort consisted of twelve males (34.3%) and twenty-three females (65.7%). The English proficiency levels among the students were distributed as follows: B1 (Intermediate): seven students (20%), B2 (Upper Intermediate): fourteen students (40%), C1 (Advanced): eleven students (31.4%), and C2 (Proficient): three students (8.6%). Three students (8.7%) had previously used VR headsets. Regarding medical conditions, two students (5.7%) reported having hypertension, four students (11.4%) reported having diabetes, none reported dyslipidemia, three students (8.6%) reported heart condition, and two students (5.7%) had a family history of cardiovascular disease

### Analysis of learning outcomes

#### Normality test

The normality tests revealed that most variables did not adhere to a normal distribution (p < 0.05). The exception was Total_mani (OSCE test grade) which showed a p-value greater than 0.05. This indicates that this variable may conform to a normal distribution. The Shapiro-Wilk test indicated non-normality (p < 0.05) for Total_VR, but the Kolmogorov-Smirnov test results were inconclusive.

#### Wilcoxon Signed-Rank Test (Table 1)

The Wilcoxon Signed-Rank test was used to assess differences between the two scenarios due to the non-normal distribution of the learning variables. The mannequin scenario showed higher results than the VR scenario in pulse oximeter placement, handling, asepsis, and drainage placement. The observed variations can be ascribed to the challenge of reproducing intricate physical actions in the virtual environment, as participants relied on the VR system controller rather than their own fingers for performing operations.

**Table 1.** Wilcoxon Signed-Rank Test Results with Effect Sizes.

| Learning Item | Z | p | r |
| --- | --- | --- | --- |
| Speed_VR - Speed_mani | -0.25 | 0.806 | -0.04 |
| Pulsi_VR - Pulsi_mani | -3.00 | 0.003 | -0.50 |
| Satu_VR - Satu_mani | -1.00 | 0.316 | -0.17 |
| BP_VR - BP_mani | -1.11 | 0.268 | -0.18 |
| Electrod_VR - Electrod_mani | -1.54 | 0.124 | -0.26 |
| Handling_VR - Handling_mani | -2.56 | 0.011 | -0.43 |
| Gloves_VR - Gloves_mani | -0.34 | 0.731 | -0.06 |
| Asepsis_VR - Asepsis_mani | -2.31 | 0.021 | -0.38 |
| Drape_VR - Drape_mani | -1.14 | 0.256 | -0.19 |
| Locate_VR - Locate_mani | -0.26 | 0.796 | -0.04 |
| Angle_VR - Angle_mani | -1.27 | 0.203 | -0.21 |
| Gguide_VR - Guide_mani | -1.74 | 0.083 | -0.29 |
| Drainage_VR - Drainage_mani | -2.34 | 0.019 | -0.39 |
| Total_VR -Total_mani | -0.58 | 0.561 | -0.10 |

The effect sizes for these comparisons were as follows: Pulsi_VR - Pulsi_mani: r = −0.500 (indicating a large effect size); the correlation between Handling_VR and Handling_mani is −0.426, indicating a medium influence; Asepsis_VR and Asepsis_mani have a negative correlation with a coefficient of −0.384, indicating a medium effect size; finally, the correlation between Drainage_VR and Drainage_mani is −0.390, indicating a medium influence.

These effect sizes demonstrate that the observed differences are not only statistically significant but also have practical significance, especially for pulse oximeter placement. There were no notable disparities observed for other items, and the magnitudes of the effects were minimal.

### Analysis of stress using biometric measurements

#### Normality test

Many variables had a non-normal distribution, as revealed by the Kolmogorov-Smirnov and Shapiro-Wilk tests (p < 0.05). The exceptions were SD1_SD2_MANI and SD1_SD2_BASAL, both of which indicated normality.

#### Friedman Test (Tables 2 and 3)

Given that the Kolmogorov-Smirnov test indicated that none of the biometric variables followed a normal distribution, the Friedman Test was employed to analyze whether there were differences in the biometric parameters among the three scenarios: basal, mannequin, and virtual reality.

- **LF and HF**: There were no significant differences in LF and HF values across the three scenarios (Chi-square = 2.643, df = 2, p = 0.267; and Chi-square = 2.951, df = 2, p = 0.229, respectively). LF/HF: there were no significant differences in LF/HF ratio and rMSSD values across the three conditions as well (Chi-square = 4.133, df = 2, p = 0.127; Chi-square = 3.211, df = 2, p = 0.201, respectively).
- **PNN20**: There were significant differences in pNN20 values across the three conditions (Chi-square = 7.393, df = 2, p = 0.025), with higher values observed in the basal state compared to VR and mannequin scenarios. Similar results were found with PNN50 (Chi-square = 9.435, df = 2, p = 0.009), with higher values observed in the basal condition compared to mannequin and VR.
- **SD1/SD2**: There were significant differences in SD1/SD2 ratio values across the three conditions (Chi-square = 14.157, df = 2, p = 0.001), with higher values in the mannequin group compared to basal and VR.

**Table 2.**
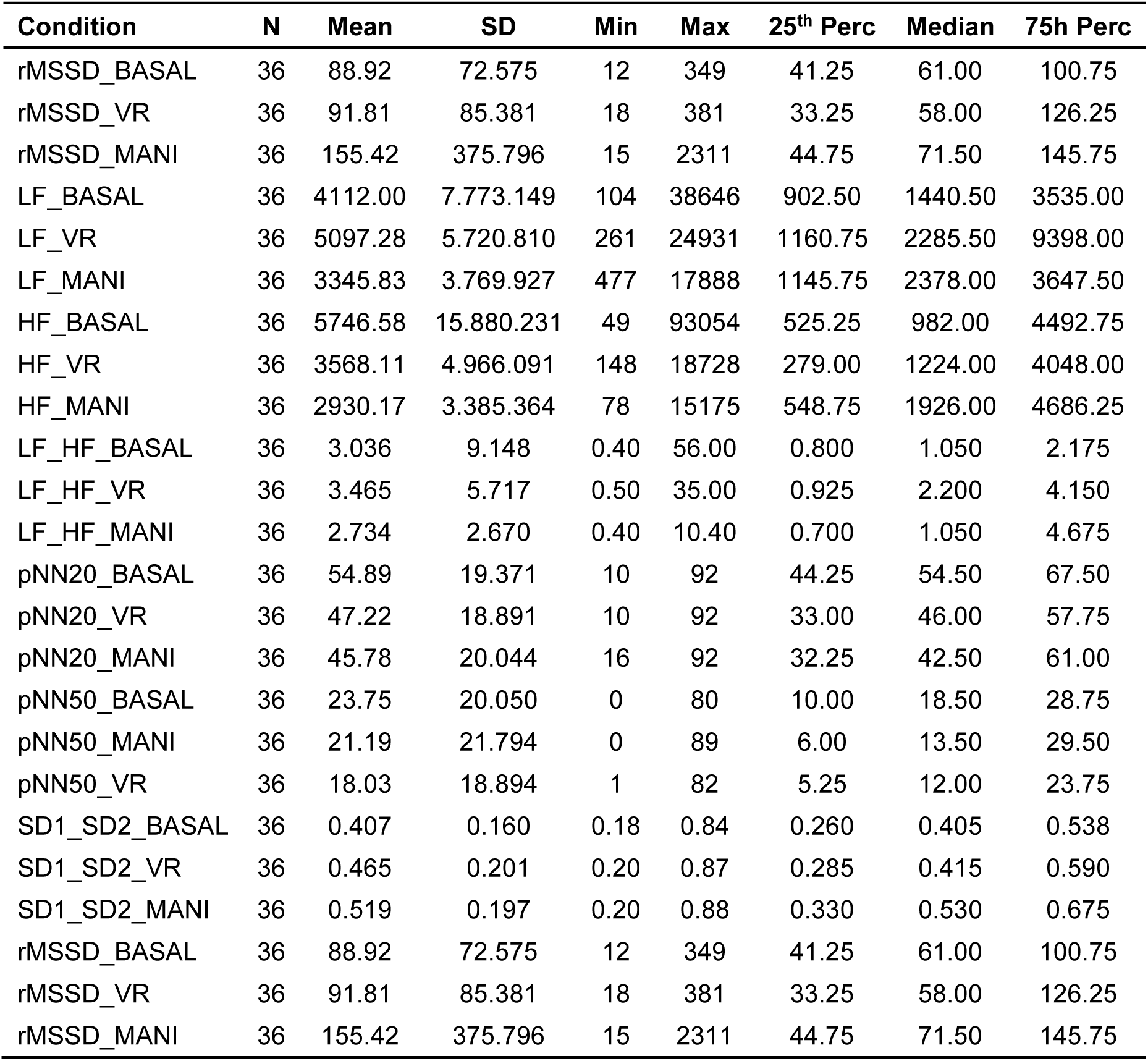
Descriptive Statistics for Biometric Parameters.

| **Condition** | **N** | **Mean** | **SD** | **Min** | **Max** | **25<sup>th</sup> Perc** | **Median** | **75h Perc** |
| --- | --- | --- | --- | --- | --- | --- | --- | --- |
| rMSSD_BASAL | 36 | 88.92 | 72.575 | 12 | 349 | 41.25 | 61.00 | 100.75 |
| rMSSD_VR | 36 | 91.81 | 85.381 | 18 | 381 | 33.25 | 58.00 | 126.25 |
| rMSSD_MANI | 36 | 155.42 | 375.796 | 15 | 2311 | 44.75 | 71.50 | 145.75 |
| LF_BASAL | 36 | 4112.00 | 7.773.149 | 104 | 38646 | 902.50 | 1440.50 | 3535.00 |
| LF_VR | 36 | 5097.28 | 5.720.810 | 261 | 24931 | 1160.75 | 2285.50 | 9398.00 |
| LF_MANI | 36 | 3345.83 | 3.769.927 | 477 | 17888 | 1145.75 | 2378.00 | 3647.50 |
| HF_BASAL | 36 | 5746.58 | 15.880.231 | 49 | 93054 | 525.25 | 982.00 | 4492.75 |
| HF_VR | 36 | 3568.11 | 4.966.091 | 148 | 18728 | 279.00 | 1224.00 | 4048.00 |
| HF_MANI | 36 | 2930.17 | 3.385.364 | 78 | 15175 | 548.75 | 1926.00 | 4686.25 |
| LF_HF_BASAL | 36 | 3.036 | 9.148 | 0.40 | 56.00 | 0.800 | 1.050 | 2.175 |
| LF_HF_VR | 36 | 3.465 | 5.717 | 0.50 | 35.00 | 0.925 | 2.200 | 4.150 |
| LF_HF_MANI | 36 | 2.734 | 2.670 | 0.40 | 10.40 | 0.700 | 1.050 | 4.675 |
| pNN20_BASAL | 36 | 54.89 | 19.371 | 10 | 92 | 44.25 | 54.50 | 67.50 |
| pNN20_VR | 36 | 47.22 | 18.891 | 10 | 92 | 33.00 | 46.00 | 57.75 |
| pNN20_MANI | 36 | 45.78 | 20.044 | 16 | 92 | 32.25 | 42.50 | 61.00 |
| pNN50_BASAL | 36 | 23.75 | 20.050 | 0 | 80 | 10.00 | 18.50 | 28.75 |
| pNN50_MANI | 36 | 21.19 | 21.794 | 0 | 89 | 6.00 | 13.50 | 29.50 |
| pNN50_VR | 36 | 18.03 | 18.894 | 1 | 82 | 5.25 | 12.00 | 23.75 |
| SD1_SD2_BASAL | 36 | 0.407 | 0.160 | 0.18 | 0.84 | 0.260 | 0.405 | 0.538 |
| SD1_SD2_VR | 36 | 0.465 | 0.201 | 0.20 | 0.87 | 0.285 | 0.415 | 0.590 |
| SD1_SD2_MANI | 36 | 0.519 | 0.197 | 0.20 | 0.88 | 0.330 | 0.530 | 0.675 |
| rMSSD_BASAL | 36 | 88.92 | 72.575 | 12 | 349 | 41.25 | 61.00 | 100.75 |
| rMSSD_VR | 36 | 91.81 | 85.381 | 18 | 381 | 33.25 | 58.00 | 126.25 |
| rMSSD_MANI | 36 | 155.42 | 375.796 | 15 | 2311 | 44.75 | 71.50 | 145.75 |

**Table 3.**
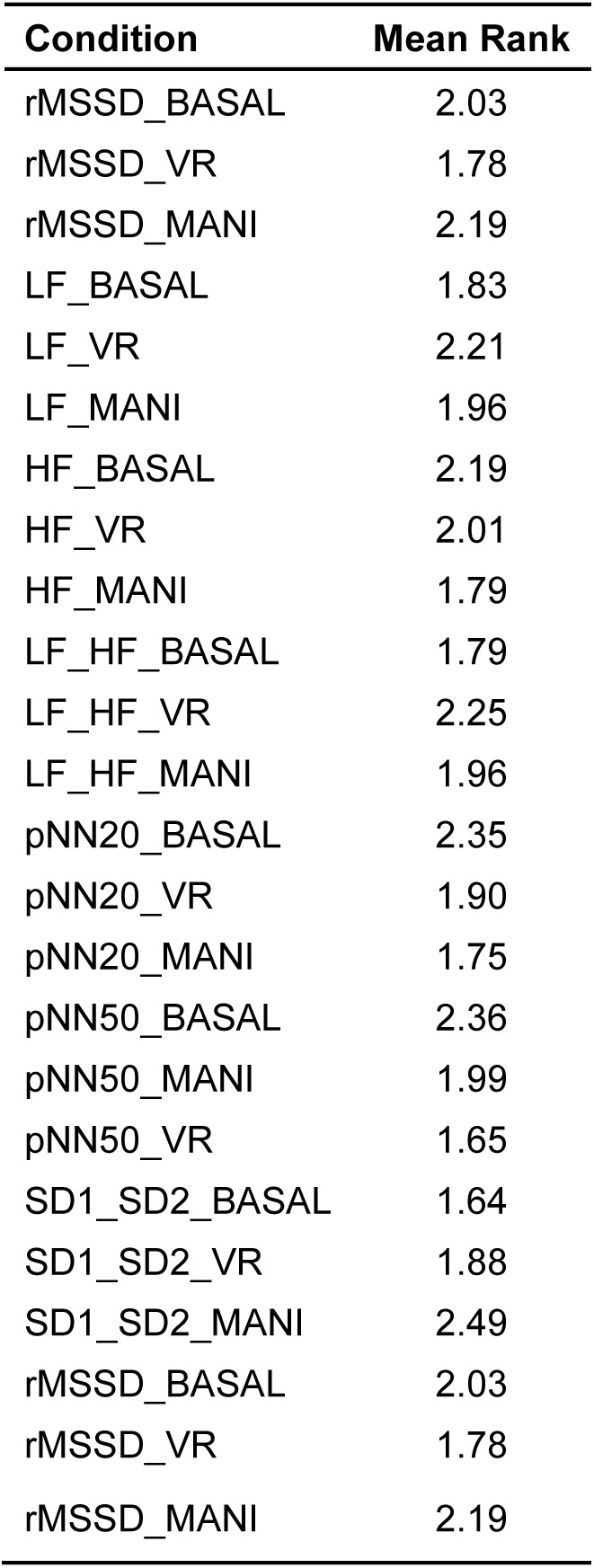
Friedman Test Results.

| Condition | Mean Rank |
| --- | --- |
| rMSSD_BASAL | 2.03 |
| rMSSD_VR | 1.78 |
| rMSSD_MANI | 2.19 |
| LF_BASAL | 1.83 |
| LF_VR | 2.21 |
| LF_MANI | 1.96 |
| HF_BASAL | 2.19 |
| HF_VR | 2.01 |
| HF_MANI | 1.79 |
| LF_HF_BASAL | 1.79 |
| LF_HF_VR | 2.25 |
| LF_HF_MANI | 1.96 |
| pNN20_BASAL | 2.35 |
| pNN20_VR | 1.90 |
| pNN20_MANI | 1.75 |
| pNN50_BASAL | 2.36 |
| pNN50_MANI | 1.99 |
| pNN50_VR | 1.65 |
| SD1_SD2_BASAL | 1.64 |
| SD1_SD2_VR | 1.88 |
| SD1_SD2_MANI | 2.49 |
| rMSSD_BASAL | 2.03 |
| rMSSD_VR | 1.78 |
| rMSSD_MANI | 2.19 |

### Self-perceived difficulty analysis

#### Normality test

The normality tests indicated that several NTLX parameters do not follow a normal distribution, specifically MENTAL_DEM_VR, PHYSICAL_DEM_MANI, PHYSICAL_DEM_VR, PERFORMANCE_VR, EFFORT_VR, FRUSTRATION_MANI, and FRUSTRATION_VR.

#### Wilcoxon Signed-Rank (Tables 4 and 5)

The Wilcoxon Test was employed to analyze differences in NTLX parameters between the mannequin and the VR scenarios. Significant differences were found in mental demand and effort, with both being lower in the VR scenario. No significant differences were observed for physical demand, temporal demand, performance, and frustration.

**Table 4.** Descriptive Statistics for NTLX Parameters.

| Variable | N | Mean | SD | Min | Max | 25<sup>th</sup> Perc | Median | 75<sup>h</sup> Perc |
| --- | --- | --- | --- | --- | --- | --- | --- | --- |
| MENTAL_DEM_MANI | 36 | 11.97 | 4.116 | 2 | 19 | 9.00 | 13.00 | 14.75 |
| PHISYCAL_DEM_MANI | 36 | 7.72 | 4.425 | 1 | 18 | 4.00 | 7.50 | 10.00 |
| TEMPO_DEM_MANI | 36 | 10.03 | 4.359 | 1 | 19 | 7.00 | 10.00 | 13.00 |
| PERFORMANCE_MANI | 36 | 8.67 | 4.414 | 1 | 18 | 5.00 | 9.00 | 11.75 |
| EFFORT_MANI | 36 | 11.61 | 3.774 | 4 | 18 | 8.25 | 12.00 | 14.00 |
| FRUSTRATION_MANI | 36 | 7.89 | 4.961 | 1 | 17 | 3.00 | 7.50 | 12.00 |
| MENTAL_DEM_VR | 62 | 10.13 | 4.611 | 1 | 20 | 5.75 | 10.50 | 14.00 |
| PHISYCAL_DEM_VR | 62 | 6.18 | 4.615 | 1 | 18 | 3.00 | 4.00 | 8.25 |
| TEMPO_DEM_VR | 62 | 9.82 | 4.091 | 1 | 18 | 7.00 | 10.00 | 12.25 |
| PERFORMANCE_VR | 62 | 7.50 | 4.218 | 3 | 17 | 4.00 | 6.00 | 11.00 |
| EFFORT_VR | 62 | 9.63 | 4.553 | 1 | 18 | 5.75 | 9.00 | 13.00 |
| FRUSTRATION_VR | 62 | 6.74 | 4.637 | 1 | 18 | 2.75 | 5.00 | 11.00 |

**Table 5.** Wilcoxon Signed-Rank Test Results.

| Item | N Neg<br>Rank | Mean<br>Rank | Sum of<br>Ranks | N Post<br>Ranks | Mean<br>Rank | Sum of<br>Ranks | Ties | Z | p |
| --- | --- | --- | --- | --- | --- | --- | --- | --- | --- |
| MENTAL_DEM_MANI | 21 | 19.07 | 400.50 | 12 | 13.38 | 160.50 | 3 | -2.147 | 0.032 |
| PHISYCAL_DEM_MANI | 22 | 17.82 | 392.00 | 13 | 18.31 | 238.00 | 1 | -1.263 | 0.207 |
| TEMPO_DEM_MANI | 20 | 17.20 | 344.00 | 15 | 19.07 | 286.00 | 1 | -0.476 | 0.634 |
| PERFORMANCE_MANI | 17 | 19.97 | 339.50 | 16 | 13.84 | 221.50 | 3 | -1.057 | 0.290 |
| EFFORT_MANI | 23 | 18.91 | 435.00 | 11 | 14.55 | 160.00 | 2 | -2.356 | 0.018 |
| FRUSTRATION_MANI | 19 | 15.29 | 290.50 | 12 | 17.13 | 205.50 | 5 | -0.834 | 0.404 |
| MENTAL_DEM_VR | 21 | 19.07 | 400.50 | 12 | 13.38 | 160.50 | 3 | -2.147 | 0.032 |
| PHISYCAL_DEM_VR | 22 | 17.82 | 392.00 | 13 | 18.31 | 238.00 | 1 | -1.263 | 0.207 |
| TEMPO_DEM_VR | 20 | 17.20 | 344.00 | 15 | 19.07 | 286.00 | 1 | -0.476 | 0.634 |
| PERFORMANCE_VR | 17 | 19.97 | 339.50 | 16 | 13.84 | 221.50 | 3 | -1.057 | 0.290 |
| EFFORT_VR | 23 | 18.91 | 435.00 | 11 | 14.55 | 160.00 | 2 | -2.356 | 0.018 |
| FRUSTRATION_VR | 19 | 15.29 | 290.50 | 12 | 17.13 | 205.50 | 5 | -0.834 | 0.404 |

## Discussion

The findings of this study demonstrate disparities in learning achievements between the conventional mannequin and virtual reality scenarios, specifically in the areas of pulse oximeter placement, handling, asepsis, and drainage placement. The mannequin model yielded superior scores in these tasks. Regarding biometric stress analysis, significant variations were observed in the time and non-linear characteristics of heart rate variability among the three conditions (basal, VR, mannequin). Higher values were observed in the basal state for pNN20 and pNN50, and for the SD1/SD2 ratio in the mannequin scenario. Furthermore, the examination of self-perceived difficulty revealed that participants reported experiencing less mental strain and exerting less effort in the VR scenario compared to the mannequin scenario.

### Learning Outcomes

The superior performance reported in the mannequin scenario for tasks such as pulse oximeter insertion, handling, asepsis, and drainage placement can be attributed to the haptic input and lifelike nature offered by actual mannequins, which are difficult to reproduce in virtual reality environments. This discovery is consistent with other research indicating that fine motor abilities are more effectively developed through physical feedback [15]. The utilization of virtual reality (VR) controllers has a detrimental effect on performance because of the absence of tactile feedback.

The use of 3D modeling in medical education represents a notable progress. Interactive 3D modeling technologies in cardiovascular surgery teaching exemplify their capacity for transformation. In their study, Gerrah & Haller [16] demonstrated the efficacy of utilizing Meshmixer to manipulate patient-specific anatomical models in real-time, thereby improving the comprehension of intricate cardiovascular operations for both patients and doctors. Open-source software has also demonstrated its efficacy in generating intricate 3D organ models, providing a cost-efficient substitute for proprietary software. In a study conducted by Cross [17], it was shown that 3D Slicer, Meshmixer, and Blender may be effectively utilized to create and manipulate cardiac models and LVAD placements. The study highlighted the practicality and ease of use of these tools in clinical settings. The development of a 3D-printed mannequin for pericardiocentesis training required sophisticated modeling and practical design, resulting in a lifelike and highly instructive model that accurately represents anatomical features.

The utilization of virtual reality (VR) in intensive care training caters to the requirement for specialized instruction in advanced medical practices. According to Lerner [18], VR technology can replicate intricate medical situations, offering a secure setting for trainees to improve their abilities without actual dangers. Although virtual reality (VR) has its limits in replicating subtle tactile sensations, both 3D-printed models and VR models successfully taught important skills for pericardiocentesis, without any notable disparities in overall learning results. The immersive quality of virtual reality (VR) improves the acquisition and transfer of skills, increasing trainees’ interest and motivation. This is particularly important in intensive care training, as demonstrated by Kyaw’s analysis in 2019[19].

Immersive virtual reality (VR) stations have been developed for Objective and Structured Clinical Examination (OSCE), and the adaptation of OSCE to a virtual platform highlights the potential of VR in medical education [20]. Our preliminary investigation revealed that the effectiveness of learning was similar when using virtual reality (VR) and mannequin models, except for tasks that involved precise control of small muscle movements. The limitation mentioned is a direct result of the current level of virtual reality (VR) technology. However, continuous research efforts are focused on enhancing haptic feedback.

### Biometric Stress Analysis

The notable results in pNN20, pNN50, and SD1/SD2 ratios indicate that the lifelikeness of mannequins produces a distinct stress reaction in contrast to virtual reality. The findings of this study are consistent with previous research conducted by Hernández-Mustieles [21] on the physiological stress reactions related to biometrics in educational environments. Using HRV analysis in VR simulations, especially in medical education, offers an original approach for assessing stress levels and physiological reactions. Aganov’s study in 2020[22] indicated that VR interventions that have been altered can have a notable impact on short-term heart rate variability and degrees of perceived anxiety. This discovery has significant importance in the field of medical education, as the ability to effectively manage stress and anxiety is essential for effective learning and performance.

Acquiring the skill to manage stress in critical medical scenarios is crucial for healthcare professionals, since it can affect their capacity to function effectively in high-pressure contexts. Training in effective stress management can result in enhanced decision-making, higher procedure accuracy, and superior patient outcomes in real-life situations. By integrating stress-inducing simulations, both virtual reality and mannequin-based training can assist students in cultivating coping strategies and resilience that are crucial in clinical practice.

The study utilized HRV as a measurement and discovered no statistically significant variations between the two models in various areas. This indicates that the virtual reality model is successful in reproducing a high-stress training environment. These findings may have substantial ramifications for the development of simulation-based education, highlighting the significance of stress management and learning efficiency.

### Self-Perceived Difficulty Interpretation

The lower mental demand and effort reported in the VR scenario compared to the mannequin scenario can be attributed to the intuitive and engaging nature of VR technology. The self-perceived difficulty was assessed using the NASA Task Load Index (NASA-TLX) in its raw form, which includes measures of mental demand, physical demand, temporal demand, performance, effort, and frustration.

Our findings indicated significant differences in mental demand and effort, with both being lower in the VR scenario. This suggests that VR simulations may reduce cognitive load and increase user engagement, making the training experience less mentally taxing. These results are consistent with previous studies that have utilized the NASA-TLX in its raw form to assess workload in VR environments. For example, Rizzo found that VR-based training for military personnel significantly reduced perceived mental workload compared to traditional methods [23]. Similarly, a study by Nayar demonstrated lower mental demand and effort in VR simulations for surgical training compared to conventional training methods [24].

However, no significant differences were found in physical demand, temporal demand, performance, and frustration between the two scenarios. This consistency across most dimensions of the NASA-TLX indicates that while VR can ease cognitive strain, it does not necessarily impact other aspects of perceived workload in the same way. The lack of significant differences in physical and temporal demands might be due to the limitations of current VR technology, such as less precise tactile feedback and the need for physical controllers.

Despite these limitations, VR has the potential to enhance the learning experience by reducing mental effort and improving engagement and motivation. Improvements in VR technology, particularly in haptic feedback, are necessary to address the limitations in fine motor skill replication and overall user experience. These enhancements could further reduce self-perceived difficulty and enhance the effectiveness of VR as a training tool.

### Implications for Training

The results of this study have important consequences for medical education and simulation training, showing that pericardiocentesis abilities may be efficiently learned utilizing either standard mannequin models or virtual reality systems. This provides empirical evidence, confirming that virtual reality is an inventive and economical approach for acquiring proficiency in intricate medical procedures. Traditional mannequins are highly effective for developing fine motor skills because they provide tactile input and are very realistic. These models also equip students with the necessary skills to manage the pressures of actual medical procedures, as demonstrated by the notable physiological stress reactions seen.

In contrast, virtual reality simulations decrease the mental burden and exertion, hence improving cognitive involvement and user motivation. VR is especially advantageous for the initial stages of learning or for training situations that include repetitive and intricate tasks, as it reduces cognitive burden. Moreover, virtual reality (VR) offers a flexible and economical method for medical training, particularly in places with limited resources or remote locations without access to sophisticated training facilities [25]. Due to its low production cost and scalability, VR technology enables a comprehensive training experience that benefits not only medical students and nurses, but also experienced professionals like residents and consultants.

The integration of 3D printed traditional mannequins with VR simulations provides a holistic training method that maximizes learning outcomes by leveraging the advantages of each modality. Training programs can use virtual reality technology to engage learners cognitively at the beginning, and then use traditional mannequins to perfect their skills. This combination improves both the efficiency of learning and the retention of acquired skills. Advancements in virtual reality (VR) technology, namely in haptic feedback, will boost its ability to accurately recreate the tactile sensations required for the development of precise motor skills.

Furthermore, the adaptability of 3D models goes beyond this model and can be applied to many invasive procedures such as thoracentesis, paracentesis, and central venous catheterization. The capacity to adapt allows for a wide range of training scenarios, making these models highly effective in various medical teaching environments. The versatility of VR across several platforms boosts its practicality, facilitating its integration into diverse educational settings.

In summary, the findings of this study help to promote the accessibility of sophisticated clinical simulation, especially in limited environments. Medical educators can enhance the effectiveness and comprehensiveness of training by combining the strengths of traditional mannequins and virtual reality (VR) simulations. This approach improves patient care and safety.

### Limitations

It is important to consider the limitations of this study while analyzing the findings. The study’s non-randomized design may create selection bias, which could restrict the applicability of the findings. Subsequent investigations should utilize randomized controlled trials to yield more substantial data.

The lack of randomization in the sequence of scenarios (mannequin vs. VR) that students encountered may result in order effects. The order in which individuals underwent the instruction could impact their performance and perceptions, thereby introducing a confounding factor into the results. To address this problem, future studies should employ randomization in the sequence of events.

Furthermore, the research was conducted exclusively at a single facility in a nation with an elevated level of income, thus impacting the generalizability of the findings. The findings may not be readily transferable to other contexts, especially in low- to middle-income nations where resources and training circumstances can vary considerably.

In addition, the study’s statistical power and ability to identify lower effect sizes may be restricted due to the small sample size of only 35 participants (n=35). Further research involving larger sample sizes and more diverse participant groups is required to validate and expand upon these findings.

Although the study acknowledged the participants’ familiarity with VR technology, only 8.7% of them had previous experience with VR headgear. This small percentage could potentially impact the overall findings. Future research should aim to include a more diverse sample in terms of virtual reality (VR) experience to gain a more comprehensive understanding of how it affects training outcomes.

The study also utilized self-reported measures of difficulty using the NASA Task Load Index, which, although validated, are subjective and susceptible to individual perceptions and biases. In future studies, it is advisable to incorporate objective measurements of workload and performance alongside self-reported data to enhance the accuracy and comprehensiveness of the findings.

Moreover, the present condition of virtual reality (VR) technology, which includes restrictions in haptic feedback, could impact the capacity to compare VR with mannequin-based training. Although ongoing research endeavors to enhance VR’s capacity to reproduce tactile experiences, it is important to recognize that these technological limitations currently exist.

While there are limits, this study offers useful insights on the efficacy of virtual reality (VR) and traditional mannequins in medical training. It also identifies opportunities for future research and technical advancement.

### Future Directions

The results of this study indicate numerous significant areas for further investigation in the field of medical education and simulation training. First, it is necessary to conduct larger research that involve more diverse and randomly selected populations to validate and broaden the observed findings. Utilizing randomized controlled trials can assist in reducing selection bias and yield more reliable evidence on the efficacy of VR and traditional mannequin-based training.

Longitudinal studies should investigate the enduring effects of VR and traditional mannequin training on the retention of skills and clinical performance over an extended period. Gaining an understanding of the long-term retention of skills will offer useful insights into the lasting effectiveness of various training methods.

It is essential to tackle the technological constraints of existing virtual reality (VR) systems, especially in terms of haptic feedback. Subsequent investigations should prioritize the enhancement of VR technology to imitate the tactile sensations and lifelike qualities of actual mannequins more accurately. This involves the creation of sophisticated haptic devices that provide enhanced and lifelike touch experiences.

There is a need to conduct greater research on the scalability and cost-effectiveness of virtual reality training programs, particularly in low- to middle-income nations and rural places. Exploring the potential of VR to equalize access to topnotch medical training would assist in addressing inequalities in educational resources and enhancing the availability of advanced training.

Subsequent research should also consider the influence of previous exposure to augmented reality on the results of training. By incorporating a sample that is more evenly distributed in terms of familiarity with virtual reality it is possible to ascertain the impact of prior exposure on learning and performance. This, in turn, can assist in developing training programs that are beneficial for both individuals new to this technology and those who are experienced users.

Incorporating biometric indicators, such as heart rate variability, into future studies can yield a more comprehensive understanding of physiological reactions to virtual reality and mannequin-based training. This has the potential to result in the creation of stress management programs within medical education, hence improving overall learning efficiency.

By including invasive procedures like thoracentesis, paracentesis, and central venous catheterization into augmented or virtual simulations, the range of applications for VR can be expanded. Research should assess the efficacy of this technology in these domains, developing flexible training modules that may be adjusted to various clinical proficiencies.

To summarize, future research should focus on verifying and expanding upon the results of this study by resolving its limitations and investigating innovative technology breakthroughs and applications of virtual reality in medical education. This approach will facilitate the complete utilization of virtual reality in augmenting medical training and boosting patient care.

## Conclusion

Our findings enhance the comprehension of simulation-based medical education, indicating that a synergistic strategy utilizing both conventional mannequins and virtual reality simulations could provide a more thorough and efficient teaching experience. The utilization of virtual reality for cognitive engagement and mannequins for hands-on practice in this hybrid model has the potential to optimize skill acquisition and retention.

Subsequent investigations should prioritize larger and more varied participant groups to validate these findings. Additionally, it is important to examine the long-term effects of both training methods on the retention of skills. Furthermore, efforts should be made to enhance virtual reality technology to simulate tactile sensations more accurately. Incorporating biometric data, such as heart rate variability, could improve our comprehension of physiological reactions to various training techniques and guide the creation of stress management programs in medical education.

The results of this study endorse the worldwide implementation of virtual reality technology as a valuable addition to conventional mannequin-based instruction in medical education. Through the resolution of existing technology constraints and the ongoing exploration of novel applications, virtual reality holds the capacity to augment the caliber and availability of medical training, hence enhancing patient care and safety.

By leveraging the strengths of both scenarios, educators can create a comprehensive training curriculum that addresses both cognitive and practical skill development. This approach will not only improve the proficiency of medical trainees but also ensure better preparedness for real-world clinical challenges, ultimately leading to improved patient outcomes.

## Author Contributions

ARL designed the study, collected the data, performed the statistical analysis, and wrote up the study as principal investigator. RG participated in the development and correction of the virtual scenario LZ performed the programming of the virtual scenario according to the created model ARN participated in the performance tests and in the design and collection of biometric parameters. AGP participated in the design and adaptation of the simulated scenarios. PCF participated in proofreading and supervision of the study design, statistical analysis, and supervision of the writing of the paper.

## Funding

This research received no external funding.

## Institutional Review Board Statement

The study was conducted in accordance with the Declaration of Helsinki,and approved by Ethics Committee for Research with Medicines of HM Hospitales (CEIm HM Hospitales Code: 18.12.1339-GHM, 01/16/2019).

## Informed Consent Statement

Written informed consent was obtained from all subjects involved in the study.

## Data Availability Statement

The original data presented in the study are openly available FigShare at https://doi.org/10.6084/m9.figshare.26215184.v1. Also pericardium molds for 3D printing can be downloaded from https://doi.org/10.6084/m9.figshare.26219135

## Acknowledgments

We extend our heartfelt gratitude to the dedicated specialists from the Intensive Care Unit at HM Montepríncipe Hospital who played a crucial role in the validation of both models presented in this study. We are particularly thankful to Dra. Fdez del Cabo, Dra. Ginestal, Dr. Waez, Dr. Añel, and Dr. Rodriguez for their invaluable contributions and expert insights, which have significantly enriched this work. Their commitment and meticulous diligence have been essential in ensuring the robustness and reliability of our findings.

## Conflicts of Interest

The authors declare no conflicts of interest.

## Appendix A

Links to pericardiocentesis procedure video:

- https://youtu.be/ox-2LP_3q_k?si=S93Dr3aB27IizWrr
- https://youtu.be/OGhQYUfpX2I?si=nvTU0-yv9qGT36YI

## Appendix B

Biosignal Plux HRV Analysis in one student in the two scenarios.

### Mannequin scenario

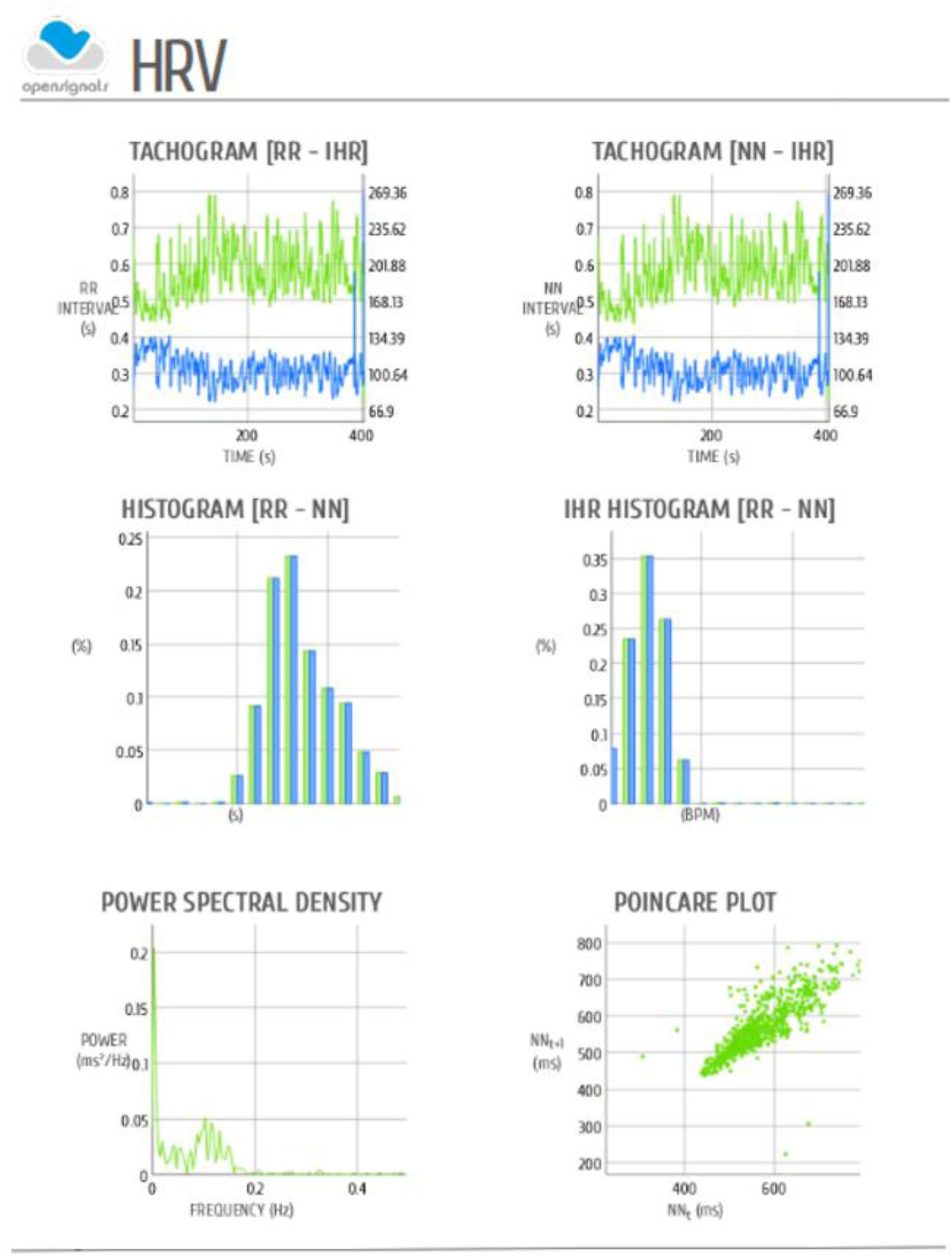

### VR scenario

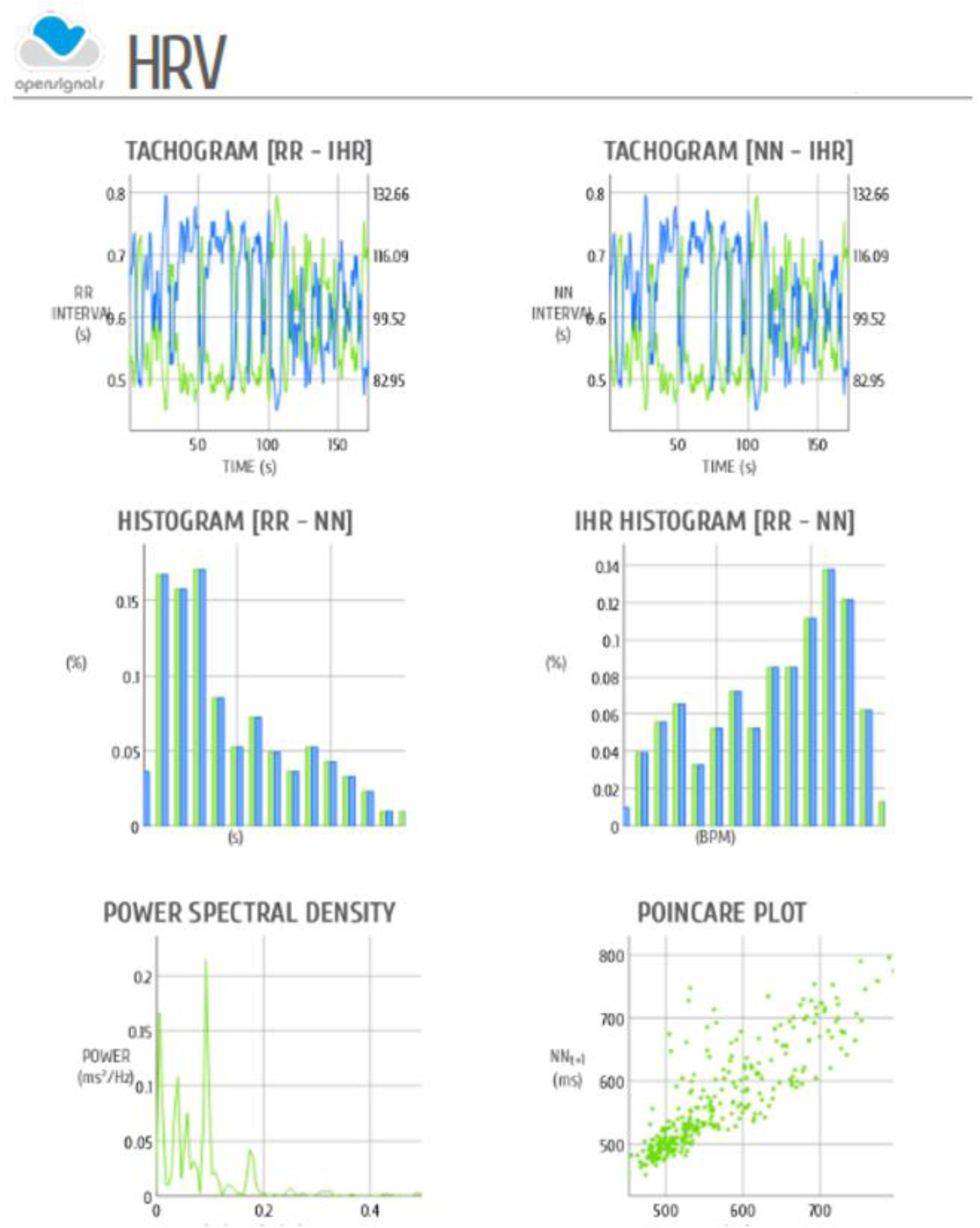

## Notes

### Competing Interest Statement

The authors have declared that no competing interests exist.

### Funding Statement

The author(s) received no specific funding for this work.

### Author Declarations

Ethics committee of HM Hospitales gave ethical approval for this work

### Summary of Updates

Revised and updated text to clarify results and discussion

